# IMPACT smoking cessation support for people with severe mental illness in South Asia (IMPACT 4S): a protocol for a randomised controlled feasibility trial of a combined behavioural and pharmacological support intervention

**DOI:** 10.1101/2021.11.03.21265856

**Authors:** Papiya Mazumdar, Gerardo Zavala, Faiza Aslam, Krishna Prasad Muliyala, Santosh Kumar Chaturvedi, Arun Kandasamy, Asad Nizami, Baha Ul Haq, Ian Kellar, Cath Jackson, Heather Thomson, David McDaid, Kamran Siddiqi, Catherine Hewitt, Najma Siddiqi, Simon Gilbody, Pratima Murthy, Noreen Mdege

## Abstract

**Introduction:** The prevalence of smoking is high among people living with severe mental illness (SMI). Evidence on feasibility, acceptability and effectiveness of smoking cessation interventions among smokers with SMI is lacking, particularly in low- and middle-income countries. We aim to test the feasibility and acceptability of delivering an evidence-based intervention (i.e., the IMPACT 4S intervention) that is a combination of behavioural support and smoking cessation pharmacotherapies among adult smokers with SMI in India and Pakistan. We will also test the feasibility and acceptability of evaluating the intervention in a randomised controlled trial.

**Methods:** We will conduct a parallel, open label, randomised controlled feasibility trial among 172 (86 in each country) adult smokers with SMI in India and Pakistan. Participants will be allocated 1:1 to either Brief Advice or the IMPACT 4S intervention. BA comprises a single five-minute BA session on stopping smoking. The IMPACT 4S intervention comprises behavioural support delivered in up to 15 one-to-one, face-to-face or audio/video, counselling sessions, with each session lasting between 15 and 40 minutes; nicotine gum and/or bupropion; and breath carbon monoxide monitoring and feedback. The outcomes are recruitment rates, reasons for ineligibility/non-participation/non-consent of participants, length of time required to achieve required sample size, retention in study and treatments, intervention fidelity during delivery, smoking cessation pharmacotherapy adherence and data completeness. A process evaluation will also be conducted.

**Ethics and dissemination:** The study has been approved by the University of York’s Health Sciences Research Governance Committee; Health Ministry Screening Committee, India; the Ethics Committee (Behavioural Sciences Division), NIMHANS, Bangalore, India; National Bioethics Committee Pakistan and; Institutional Research and Ethics Forum of Rawalpindi Medical University, Pakistan. Feasibility study results will be disseminated through peer-review articles, and presentations at national and international conferences and policy-engagement forums.

**Trial registration:** ISRCTN34399445 (Updated 22/03/2021), ISRCTN Registry https://www.isrctn.com/

## Introduction

People with severe mental illness (SMI) (schizophrenia, bipolar disorder, other psychoses and severe depressive disorder) constitute one of the world’s most vulnerable populations and face significant health and socioeconomic inequalities. Compared to the general population people with SMI have twice the rate of mortality and 10-25 years lower life expectancy.^1,2^ The drivers for these differences are coexisting physical disorders^3^ attributable to health risk behaviours, such as smoking, alcohol and illicit drug use, poor diet and physical inactivity.^4^ Global health policies and research agendas have prioritised addressing the physical health of people with SMI,^5^ with an emphasis on interventions targeting health risk behaviours.^6^

Tobacco use is particularly high in people with SMI, with overall prevalence estimates ranging from 50-90%,^7^ and that of smoking being between 40 to 60%.^7,8^ Limited data from South Asian countries suggest smoking prevalence is as high as 50% among adults with SMI^8^ which is significantly higher than in the general population (e.g., 10% in India,^9^ and 12.4% in Pakistan^10^). People with SMI also smoke more heavily, have more severe nicotine dependence, and suffer worse health outcomes as a result of tobacco use compared to those without SMI.^11^ Although smoking is the most common completely modifiable health risk behaviour,^12^ the benefits of smoking cessation programmes are yet to reach people with SMI.^12^ For example, smoking cessation counselling delivered to individuals or groups significantly increase the chances of smoking cessation in the general population,^13,14^ but results among people with SMI have been mixed.^15-17^ Moreover, the evidence among people with SMI is mostly from high income countries (HICs). HIC studies have also shown that pharmacotherapies such as bupropion and varenicline, alone^18^ or in combination with behavioural support,^19^ are safe, effective, and acceptable for smoking cessation in adults with SMI.

Tailored smoking cessation interventions that take into account some of the challenges among people with SMI have been developed for HICs,^17^ but not for LMICs settings. Significant differences in cultural and contextual patterns of tobacco use, health systems characteristics, and regulatory approaches make it difficult to directly translate evidence from HIC settings to LMIC contexts. For South Asia, interventions would need to consider the high prevalence of both smoking as well as other forms of tobacco use. We followed an iterative process of intervention adaptation to develop an evidence based smoking cessation intervention (i.e., the IMPACT 4S intervention) for adult smokers with SMI in India and Pakistan. The IMPACT 4S intervention is a combination of behavioural support and smoking cessation pharmacotherapy, and was adapted from three smoking cessation behavioural support intervention manuals ^20,21,22^ :the SCIMITAR+ intervention developed for adult smokers with SMI in the UK based on the UK’s National Centre for Smoking Cessation and Training standard treatment programme,^23, 24^ the TB & Tobacco intervention developed for TB patients in Bangladesh and Pakistan,^25^ and a tobacco cessation smart guide developed by the National Institute of Mental Health and Neuro Sciences Resource Centre for Tobacco Control Bangalore, India.^26^ The adaptation process will be reported elsewhere. In this study we aim to test the feasibility and acceptability of delivering the IMPACT 4S intervention, and of conducting a randomised controlled trial, among adult smokers with SMI attending mental health facilities in India and Pakistan.

### Specific objectives

We will address the following specific objectives:

1. To test the feasibility and acceptability of delivering the IMPACT 4S intervention for smoking cessation among adult smokers with SMI
2. To test the feasibility and acceptability of conducting a definitive randomised controlled trial to evaluate the effectiveness and cost-effectiveness of the IMPACT 4S intervention for smoking cessation among adult smokers with SMI

## Methods

### Study design

The study consists of two main components: i) a parallel open label randomised controlled feasibility trial over 12 months; and ii) an embedded process evaluation (implementation, mechanisms of impact). ^27^ (presented in Figure 1).

**Figure 1:**
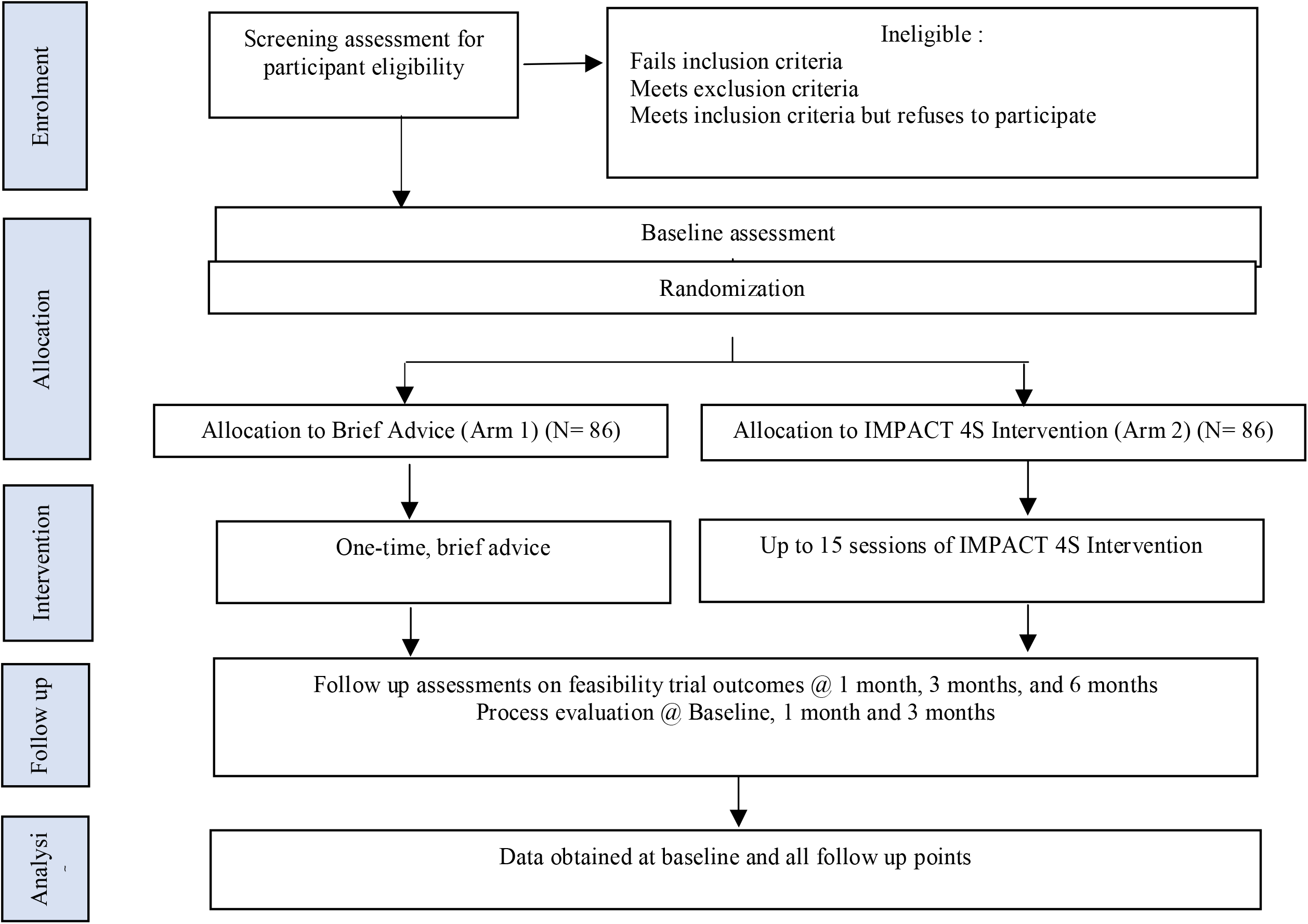
Study flow diagram depicting design of the feasibility trial and process evaluation

### Study Setting

The feasibility trial will be conducted at the National Institute of Mental Health and Neurosciences (NIMHANS), Bangalore, India, and Institute of Psychiatry (IoP), Rawalpindi, Pakistan.

### Feasibility Trial

For the parallel open label randomised controlled feasibility trial, participants will be allocated 1:1 to the following two arms:

- Brief advice (BA)
- IMPACT 4S intervention

Eligibility criteria for inclusion are: i) adults (≥18 years old) with SMI (i.e. schizophrenia, schizoaffective disorder, bipolar affective disorder, psychosis, severe depression with psychosis), ii) considered to be stable by the mental health clinical team, iii) self-reported current smoker of any form of smoked tobacco product (including cigarettes, hand rolled cigarettes/ bidis, waterpipe etc) for at least 6 months, iv) smoking on >25 days in the past month, v) clinically assessed able to provide informed consent, vi) attending participating institutions during the study period, vii) willing to quit smoking, viii) willing and able to attend up to 15 counselling sessions and ix) for India, living in urban and rural Bangalore districts or neighbouring districts and for Pakistan, living in Islamabad Capital Territory and Rawalpindi district. The exclusion criteria are: i) pregnant or breastfeeding women, and ii) the presence of comorbid drug and/or alcohol problem as this would require specialist treatment.

#### Participant recruitment and eligibility assessment

Potential participants will be informed about the study by their care team and referred to the study team if they are interested in participating. A researcher (i.e., trained psychology graduate /psychiatric social worker) will use a screening form to determine trial eligibility. Screening will be conducted either in-person or over audio/video call, according to participants’ availability and preference. All screened participants will be allocated a unique screening number, and the following anonymised individual information will be collected and stored in a secure study database: age; gender; SMI diagnosis; current smoking status; eligibility criteria; time taken for screening and reason for exclusion, where applicable. Individuals who meet the eligibility criteria will receive a detailed study information sheet (written either in English, Kannada, Telegu, Hindi and Urdu) that includes the following information: the purpose of the feasibility study; the study procedures including details of the interventions; process of randomisation; the frequency and timing of data collection; breath carbon monoxide (CO) monitoring; the potential benefits or risks of participating in the trial; information on privacy and confidentiality of the participants’ data, and how it will be processed; and participant’s rights, including the voluntary nature of the trial and the right to discontinue participation at any time without any consequences. The information sheet will also provide contact details of a research team member in case they wish to seek more information or clarity. In circumstances where a potential participant is unable to read or write, or for those recruited through an audio or video call, the researcher will read out the information sheet to both the participant and the carer if present and will respond to their queries. Participants will be given at least 24 hours, if they require, to discuss the information with family members or friends before deciding to take part.

#### Consent

Informed consent will be obtained before any study specific assessments are conducted. Once the participants are familiar with the intent and purpose of the study and have their queries addressed, those willing to participate will be invited to provide either written consent (for those screened in-person) or audio/video recorded verbal consent (for those screened over the telephone). We will follow a formal informed consent procedure for those providing written consent, where they will provide their full name and signature or, if they cannot write a thumb impression (with a witness present). One copy of the consent form will be securely stored in a locked filing cabinet at the trial sites in India and Pakistan, and the participant will get a copy of their signed informed consent form, and the information sheet to take home for their records. For few of those expected to record verbal consent through an audio or video call, the researcher will read each statement from the consent form and following their verbal consent, researchers will fill-up a written consent form for these participants to provide signature/thumb impression whenever they come at the trial site for attending sessions in-person.

Participants will be free to withdraw consent and leave the trial at any time without having to cite any reason. All participants will be provided with written or verbal contact information on who to contact, if they wish to withdraw from the trial. If a participant withdraws consent to participate, no further data will be collected from them. However, data collected up to the point of withdrawal will be retained and used in the analysis, unless there is an explicit request for withdrawal of data prior to analysis, in which case all data will be destroyed.

All consenting individuals will be entered on to a secure study database, using their name, age, address, and unique trial ID. Only the principal investigators, country investigators, trial coordinators, and researchers involved in collecting, quality checking and entering data will have access to these identifiable data at any stage of the trial, mainly to facilitate participant follow-up. Identifiable data will be stored separately from the baseline and follow-up data. For the purposes of baseline and follow-up data collection only the unique study IDs will be used, thereby ensuring anonymity of data.

#### Randomisation, trial arm allocation and blinding

Eligible participants who consent will be allocated a unique study ID and randomly assigned to one of the trial arms using a computer-generated blocked stratified (by country) randomisation sequence created using Stata version 15 (or later), with an allocation ratio of 1:1. A statistician based at the University of York, who is not involved in the recruitment of trial participants, will generate the randomisation sequence and produce opaque sealed envelopes which will be stored securely at each trial site central research office and used to randomly allocate participants to the trial arms. After a participant has provided written informed consent, a researcher will conduct a baseline assessment, then make a phone call to the country specific trial manager who will be based at a central research office. Upon being provided with basic participant information, the trial manager will allocate the next envelope in the sequence to the participant. Up until this point, both the trial manager and the researcher will be unaware of the allocation associated with the envelope. After opening the envelope, the trial manager will indicate to the researcher whether the participant will receive the IMPACT 4S intervention or brief advice and the envelope number and allocation will be recorded on the participant record.

Blinding the participants and clinicians from knowing who is receiving the IMPACT 4S intervention or BA is not feasible. Outcome data collection and data analysis will also not be fully blinded.

#### Interventions

##### Brief advice (BA)

Brief advice has been adapted from a community-based trial of tobacco cessation in India,^28^ and comprises a single, five-minute conversation on the harmful effects of tobacco and advice to stop smoking delivered by trained psychology graduates. Participants will also receive a Brief advice information leaflet containing the same advice in written format to take home.

##### IMPACT 4S Intervention

This comprises behavioural support adapted to adult smokers with SMI in India and Pakistan, breath CO monitoring and feedback, pharmacotherapy (bupropion and/or nicotine replacement therapy), and the same information leaflet as for the BA arm to take home. The intervention was co-produced in collaboration with smoking cessation specialists and behavioural scientists based in the UK, India and Pakistan, and key stakeholders (including policymakers, practitioners and adults with SMI who smoked) from India and Pakistan.

#### The behavioural support for smoking cessation

includes behaviour change techniques (BCTs) that were identified, through literature reviews and intervention adaptation workshops with stakeholders, as important for changing smoking behaviour among adults with SMI who smoke.^29^ The BCTs were chosen from a taxonomy within a Western context as no similar taxonomy has been developed for smoking cessation in LMICs^23^ and alongside BCTs previously used for smoking cessation in India.^26^ The BCTs are goal setting, problem solving, social support, information about health consequences and adding objects to the environment. Participants will be encouraged to: (1) quit abruptly, or cut-down the present level of smoking, in a managed way, until they quit, (2) set out their own quit date and (3) make several attempts to quit if their initial attempt fails. The behavioural support has been designed for either in-person, remote, or a combination of both in-person and remote sessions, to allow adaptation to practical feasibility and participants’ preferences. Remote sessions will be delivered using several platforms, routinely used for patient consultations at both institutions, including, telephone or video calls (e.g., WhatsApp, messenger, Zoom, etc.), taking into account participant preference. The behavioural support will comprise a maximum of 15 one-to-one counselling sessions over three months, each lasting between 15-40 minutes, determined by mode of delivery and participant need. Each session will be delivered by trained psychology graduates in Pakistan and psychology graduates or psychiatric social workers in India. Delivery will be facilitated through the use of an intervention manual/guide adapted for both in-person or remote delivery; a flipbook containing key messages on how to quit smoking; and the participants’ resource material (a set of seven-colour coded information sheets) that provides information for each session, including any relevant ‘homework tasks’ (e.g. smoking diary, planning for checklist for quit day and summarizing reasons for stopping) for participants to complete independently wherever possible/ with help of career, in advance of the next session. As these ‘homework tasks’ are key to the intervention adherence, the staff delivering the intervention will check for completion during subsequent session, and provide necessary help for completion if needed. In the case of in-person delivery, the manual or flipbook will help the staff delivering the intervention to deliver messages via photo images on the front of the slides facing participants, and text on the back of slides facing them. The same messages will be reiterated in posters and leaflets. This type of intervention delivery is familiar to health workers in these settings and has been found feasible and acceptable in South Asian countries including Pakistan.^25^ Remote delivery of the intervention will use two similar materials deployed for in-person sessions 1) intervention flipbooks; and 2) the participants’ resource materials. These materials will be posted out to participants or given to them in person, as required.

#### Exhaled Breath CO monitoring and feedback

Exhaled breath CO levels will be measured using Bedfont piCO™ Smokerlyzer® CO breath test monitor at every counselling session conducted in-person, and feedback will be provided in order to increase the chances of a successful quit attempt.^30,31^ At remotely delivered sessions where CO monitoring is not possible, the staff delivering the intervention will give information and feedback on the expected breath CO levels based on self-reported smoking behaviour or progress in quitting smoking.

#### Pharmacological support

Participant will be provided with a free three-months supply of nicotine gums or bupropion tablets either separately or in combination. Participants who opt for bupropion will be assessed for suitability by an in-house clinical psychiatrist (either in-person or through video consultation) and if found eligible, will be offered sustained-release bupropion: 150 mg/day for the first week followed by 300 mg/day intake until a maximum of three months. The patients receiving Bupropion are dispensed the medicines initially on weekly basis then fortnightly and then for a month. The patients who choose to use nicotine gums are provided advance stock for a week in the first month and then on monthly basis for rest of the two months. All participants will either collect medication in advance during their in-person visits to the study settings or will have it delivered to them at home by a research assistant. Participants will be provided with both verbal and written information on the correct dosage of medication, method of use, side effects of bupropion/nicotine gums, and contact details of team members for reporting and/or advice for medication side effects.

#### Intervention training

The duration of intervention training will be half a day for the brief advice intervention, and one to two days for the IMPACT 4S intervention. All staff delivering the interventions will attend training tailored for the specific intervention. During the intervention delivery period, the staff will also receive ongoing support and supervision from experienced team members.

#### Outcome measures

1. Recruitment rates: will be assessed as the number of participants eligible, consenting and randomised, out of those screened.
2. Reasons for ineligibility/non-participation/non-consent of participants.
3. Length of time required to achieve the required sample size.
4. Retention in study: will be assessed as the number of participants randomised who are successfully followed-up at six months.
5. Retention in treatment: will be reported as the number of Brief Advice and IMPACT 4S intervention sessions attended.
6. Intervention fidelity to delivery of the behavioural support within the IMPACT 4S intervention, as well as for BA, assessed as one measure of feasibility of intervention delivery.
7. Smoking cessation pharmacotherapy adherence: For those in the IMPACT 4S arm, adherence to smoking cessation pharmacotherapy will be assessed.
8. Data completeness: Data will be checked for completeness as another measure of acceptability and feasibility of data collection methods, and to identify problem areas and solutions.

The following data will be collected as per the schedule in Table 1:

**Table 1:** Baseline and follow-up assessment schedule

| Measurement | Baseline | 1 month<br>(Follow- | 3 months<br>(Follow- | 6 months<br>(Follow- |
| --- | --- | --- | --- | --- |

|  |  | up 1) | up 2) | up 3) |
| --- | --- | --- | --- | --- |
| Demographic data | X |  |  |  |
| Preference for mode of intervention delivery | X |  |  |  |
| Carers/ family members' involvement | X |  |  |  |
| Details of psychiatric medication | X | X | X | X |
| Smoking status (validated by CO exhaled breath test, wherever possible) | X | X | X | X |
| Nicotine dependence and urge to smoke | X | X | X | X |
| Quitting smoking intentions, motivations and behaviours | X | X | X | X |
| Mediators of smoking cessation | X |  | X | X |
| Health-related quality of life | X |  | X | X |
| Mental and physical health | X |  | X | X |
| Health services use | X |  | X | X |
| Physical body measurements (height; weight) | X |  | X | X |
| Smoking cessation pharmacotherapy adherence |  | X | X | X |
| Process evaluation questionnaire | X | X | X |  |
| Process evaluation interviews |  |  | X |  |

- Demographic data (baseline): The information on age and gender collected at the participant eligibility screening stage will be verified by asking the participant. We will also collect data on education, occupation, marital status, and on the possession of household items to determine socio-economic status following the demographics module of the WHO STEPwise Approach to NCD Risk Factor Surveillance (STEPS) questionnaire.^32^
- Psychiatric medication use (baseline, one, three and six months): Details of psychiatric medication that the participant is taking will be taken at baseline, and verified at each follow-up visit, in-order to capture any changes.
- Smoking status (baseline, one, three and six months): Self-reported or family/carer reported smoking status will be assessed using the smoking questions from the GATS survey.^33^ At six months, self-reported smoking status will be biochemically verified by a CO level of <7ppm for those reporting that they have quit smoking, as per the Russell Standard.^34^ When a participant self-reports abstinence with an elevated CO level of >7ppm, the biochemical verification will supersede the continuous and point abstinence self-report and the participant will be defined as a smoker.
- Nicotine dependence and urge to smoke (baseline, one, three and six months): The heavy smoking index will be used to assess nicotine dependence from smoked tobacco.^35^ The Mood and Physical Symptoms Scale (MPSS)^36^ will also be administered. The scale assesses withdrawal symptoms including anxiety, depression, irritability, restlessness, hunger, concentration and sleep.
- Quitting smoking intentions, motivations, and behaviours (baseline, one, three and six months): The four-item motivation to quit (MTQ) questionnaire^37,38,39^ will be used to measure motivation to quit smoking. It is scored by summing the responses to each item. The scores range from 4 to 19 where a higher score indicates greater motivation to quit. Questions from the Global Adult Tobacco Survey (GATS)^33^ will also be used to assess smoking cessation intentions, motivations, and behaviours.
- Mediators of smoking cessation (baseline, three and six months): A set of questions related to the IMPACT 4S interventions’ proposed mechanisms of action 23-25 have been developed during the intervention adaptation phase. These are beliefs about capabilities, behavioural cueing, beliefs about consequences, environmental context and resources.
- Quality of life (baseline, three and six months): The EQ-5D-5L^40,41^ will be used to measure health-related quality of life (HRQoL). All necessary permissions for the use of English and Urdu versions of EQ-5D-5L (both paper self-administered and telephone) are obtained for the period between 2019 to 2022. EQ-5D is a standardised measure of health status developed by the EuroQol Group in order to provide a simple, generic measure of health for clinical and economic appraisal, where health is characterised on five dimensions (mobility, self-care, ability to undertake usual activities, pain/discomfort, anxiety/ depression).
- Health services use (baseline, three and six months): Participant use of health services and other smoking cessation services outside the study will be assessed. A health service utilisation questionnaire previously used in some of our studies such as the MCLASS trials^42^ or the Client Service Receipt Inventory, adapted to the context of Pakistan and India, will be used to collect number and type of contacts with doctors, hospital admissions, pharmacy visits and medication prescriptions for all participants. Information on contact with traditional healers will also be recorded. We will also collect costs of intervention delivery at both trial sites.
- Mental and physical health (baseline, three and six months): PHQ-9^43^ will be used to measure depressive symptoms. This nine-item questionnaire is scored from 0 to 27, and a higher score indicates more severe depressive symptoms. The GAD-7^44^ will be used for measuring anxiety. This seven-item instrument is scored from 0 to 21, with a higher score indicating more severe anxiety. The SF-12 which consists of two subscales: a physical health component and a mental health component will also be administered. Both components are scored from 0 to 100, with 0 indicating the lowest level of health and 100 the highest level of health measured by the scale.^45^
- Physical body measurements (baseline, three and six months): Physical body measurements will include height and weight which will be measured by trained personnel according to the WHO protocols.^46^ These will be used to determine the body mass index. Physical body measurements of the trial participants will not be measured if the recruitment and assessments are conducted remotely, or the intervention is delivered remotely.
- Smoking cessation pharmacotherapy adherence (one and three months): For the IMPACT 4S group, self-reported medication adherence to smoking cessation pharmacotherapies will be assessed retrospectively based on a 4-day recall.^47^

An adherence index will be calculated by the formula:

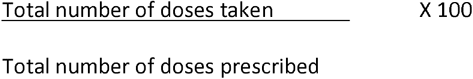

Participants with more than 80% of adherence will be considered as having high adherence and those with less than 80% will be considered as having low adherence.^48^ Participants who report missed doses will be asked to provide reasons for missing their medications.

#### Data collection

Data will be collected from participants and carers either in-person or remotely at baseline, one, three- and six-months post-randomisation. Data will either be recorded on paper format first or directly into an online survey tool (Qualtrics) using tablets^49^. Data will be collected by researchers who would have received three days of training on trial procedures including taking informed consent, administering and completing the questionnaires and ethical issues such as autonomy of individual participants on making decisions about participation, freedom to withdraw from the trial without giving any reason or consequence, privacy, confidentiality, anonymity.

#### Sample size and statistical analysis

The sample size calculations are based on estimating participation and attrition rates and event rates as the primary outcomes. If we identify 344 eligible subjects we will be able to estimate a participation rate of 50% to within a margin of error of ±5% and an attrition rate of 30% to within ±8%.^50^ Furthermore, an external feasibility study of at least 120 measured participants will provide robust estimates of event rates to inform the sample size calculation for the subsequent larger definitive fully powered trial.^51^ We will therefore aim to enrol 172 (86 per country) participants in the study in order to retain 120 for analysis assuming an attrition rate of 30%.

The following outcomes will be calculated: number of eligible patients; proportion of eligible patients approached for consent; proportion of eligible patients not approached and reasons why; proportion of patients approached who provide consent; proportion of patients approached who do not provide consent; proportion of patients providing consent who are randomised; proportion of patients randomised who do not receive the randomly allocated treatment; proportion of patients dropping out between randomisation and follow-up. A CONSORT diagram will be provided to display the flow of participants through the study. Data from the clinical outcome measures will be summarised descriptively: continuous data using mean, standard deviation, median and 25th and 75th percentiles; categorical data using number of events and percentages. The recruitment rate and 95% confidence interval (CI) will be estimated. Completion rates of all the outcome measures will be reported by study arm.

#### Analysis of economic and quality of life data

The completeness of returned HRQoL and health care utilisation data will be assessed to inform an economic evaluation alongside a full RCT. Returned service use data will be used to revise questionnaires used in a full trial. Costs of delivering the interventions, measured by recording the resources utilised in the delivery of the IMPACT 4S intervention (including behavioural support, CO monitoring and feedback, pharmacotherapy, costs for telephone/remote modes, client information-sheets, staff resource for clients’ records and information leaflet costs) and BA (including the counselling and information leaflet), will be summarised. We will also produce summaries of: the costs of intervention setup, training, as well as ongoing support and supervision of the personnel delivering the intervention for each trial arms; and the costs for participant and carer/family member’s travel costs and travel time to attend intervention. Pilot economic analysis will be conducted to inform a future economic evaluation embedded within a definitive large-scale RCT.

### Process evaluation

The process evaluation will be informed by the Medical Research Council guidance for process evaluation^27^ which identifies three components: *implementation, mechanisms of impact and context*.

#### Implementation (feasibility) and context

Once delivery of the IMPACT 4S and brief advice interventions are completed, the psychology graduates or psychiatric social workers who delivered these interventions will be interviewed to explore their experiences, including the acceptability of the interventions and the barriers and drivers to delivery including contextual factors such as the health facility environment.

#### Implementation (fidelity)

Fidelity to delivering the IMPACT 4S and interventions will be assessed using a fidelity index. This consists of two sub-indices: The Adherence Index that assesses adherence to the intervention activities; and the Quality Index, that assesses competence with which the intervention was delivered. Both indices are scored on a three-point Likert scale (0=not implemented, 1=partially implemented and 2=fully implemented).

All counselling sessions will be audio-recorded. Fidelity checks will be conducted on the sessions for 12/43 participants per trial arm, per country. Participants will be purposively selected to ensure we review fidelity for all the counsellors at different points in the study (early, middle, late).

#### Mechanisms of impact (acceptability)

To capture the views and experiences of participants, all participants will complete a short questionnaire at baseline, one and three month follow up. At baseline participants will be asked about their preferred mode for receiving the intervention and on accompanying family/ carer for the session attendance. Then, depending on their trial arm allocation, questions will explore which components of the brief advice / IMPACT 4S intervention they have engaged with, accompanying person during session attendance both in case of in-person or remote modes, the acceptability of each session including the CO monitoring, their use of pharmacotherapies and perceived impact of the interventions on their smoking behaviour. Participants in both trial arms will complete some acceptability questions on trial processes.

At three month follow-up a purposive sample of participants in each country (12-16 in the IMPACT 4S arm, 4-8 in the BA arm), reflecting a mix of men and women who have/have not quit smoking will be interviewed to collect more in-depth feedback on the topics explored in the short questionnaire.

All interviews will be conducted face-to-face or remotely, using a topic guide, and will be digitally audio-recorded. They will be between 30 to 60 minutes duration. A hermeneutics approach, which encourages participants to discuss features of the intervention to elicit data on their experience and evaluation of its delivery/receipt will be used.^52^

Participants will provide written informed consent or will provide audio-recorded consent for these interviews, separate from the trial, after having received enough information for this study component and their questions being answered. The consent procedures will be the same as for the trial.

The quantitative data from the fidelity index and questionnaire will be analysed using descriptive statistics, including means and standard deviation for continuous variables, and absolute and relative frequencies for categorical variables. Interviews will be transcribed verbatim and translated into English and analysed using the Framework Approach^53^ which is designed to address applied programme and policy-related questions. NVivo 11 software will aid data handling. Integration of interview findings with respective questionnaire data will be done using a ‘triangulation protocol’.^54^

### Adverse event (AE) monitoring

AEs will be assessed at each counselling session irrespective of the mode of delivery as well as at follow-up visits. Any potential association with study interventions or procedures will be ascertained. The researcher conducting the counselling session or follow-up visit assessments will record all directly observed AEs and all AEs reported by participants. An AE review checklist will be used at each session to explicitly prompt for symptoms relating to possible nicotine replacement therapy or bupropion related toxicities. This way we will be able to address any AEs or concerns that the participants might have. Appendix 1 (safety consideration) provides further information on expected AEs monitoring and management procedures to be followed by the study.

### Data Management Plans

#### Data storage and archiving

Study information (including consent forms, screening and enrolment data, baseline and follow-up data, audio-recordings and transcripts) will only be accessed by researchers, and will not be released without written permission, except as necessary for monitoring by study monitors or regulatory bodies, for example if the study is audited. Encrypted data transfer between the study sites and the University of York will be via secure Drop-off services available through the University of York. At the end of the study, data will be securely archived by the University of York, NIMHANS and IoP, as appropriate, for 10 years. The staff involved in the trial will be trained on data protection processes. The staff will be strictly monitored to ensure compliance with privacy standard.

#### Data monitoring

Data will be monitored for quality and completeness by a delegated researcher at the study sites, followed by a second check by the study Research Fellow at the University of York using verification, validation and checking processes. Missing data will be pursued until study end unless it causes any distress to the participant contacted. Data will be reported to the Independent Trial Steering Committee (ITSC) and Data Monitoring and Ethics Committee (DMEC) as required.

#### Ethics and other approvals

Ethics and other relevant approvals on original protocol have been sought from the University of York’s Health Sciences Research Governance Committee (Approval number: HSRGC/2019/346/D, on July 5, 2019), relevant national and institutional ethics committees (i.e., Health Ministry Screening Committee, India (Approval number: 2019-7975 on February 19, 2020) ; Ethics Committee (Behavioural Sciences Division), NIMHANS, Bangalore, India ; National Bioethics Committee Pakistan (Approval number: NBC-434 on December 10, 2019); Institutional Research and Ethics Forum of Rawalpindi Medical University (Approval number: R-48/RMU on August 24, 2019), Pakistan and from each administrative site in the participating countries prior to commencement of research activities. The approvals on amendments adjusting to the COVID-19 pandemic situation have been provided by all of the above-mentioned ethics-review boards between April-July, 2021.

#### Feasibility trial timeline

Recruitment commenced in April 2021 and shall continue until September 2021.

## Discussion

There has been almost no reduction in smoking prevalence among adult smokers with SMI, even in HICs, despite the dramatic reduction in smoking rates in the general population that has occurred during the past 40 years.^12^ The reasons for this include “diagnostic overshadowing” whereby physical health is neglected in the presence of a mental illness diagnosis; and the unsuitability of interventions designed for the general population that are predicated on high levels of cognitive function, literacy, health literacy, motivation and self-efficacy, all of which may be compromised by SMI.^4,55^ To the best of our knowledge, this will be the first randomised controlled feasibility trial to address this evidence gap in the South Asian context. We will contribute to addressing the uncertainty about the feasibility and acceptability of delivering smoking cessation interventions, and the ability to conduct smoking cessation trials, among adult smokers with SMI in LMICs. The study evaluates, for the first time, the feasibility and acceptability of an innovative, bespoke, and culturally adapted IMPACT 4S intervention that combines behavioural support and pharmacotherapy, and can be delivered either in-person, remotely, or as a hybrid of in-person and remote delivery modes among adult smokers with SMI in India and Pakistan who smoke. If the IMPACT 4S intervention is found to be feasible and acceptable to deliver and evaluate in a definitive trial, then we will seek funding for a full trial to generate robust evidence on its effectiveness and cost-effectiveness for smoking cessation among adult smokers with SMI in the South Asian context.

The nature of the interventions being evaluated does not allow blinding of participants, or the staff delivering the interventions, to the allocation. In addition, data analysis will not be blinded. Nevertheless, the study has a number of strengths, including random allocation of participants to the study arms, allocation concealment, and the collection of a comprehensive list of data variables that are likely to be collected in a full trial. In addition, the process evaluation will allow collection of qualitative feasibility and acceptability data from the perspective of both the participants and the staff delivering the interventions; including data on the different intervention components.

The results of the feasibility trial will be published in peer-reviewed journals and presented at relevant national and international conferences, workshops, and policy-engagement forums. Authorship credit shall be based on significant contribution to conceptual design and structure, execution of analysis and interpretation of data, and drafting, revision and critical review of intellectual contents and final approval of the manuscripts.

### Potential amendments

The study may be stopped if, guided by the Independent Trial Steering Committee (ITSC) and Data Monitoring and Ethics Committee (DMEC), new literature indicates definitive findings in terms of benefit or side effects of the intervention components (i.e., the behavioural support and/or the pharmacological components). The reporting of AEs might also indicate the need for study protocol review in terms of medication or rescue medicine usage. Considering the present Covid-19 pandemic and related uncertainties, the trial might need to revise the 6 month follow-up assessment. The decision on revising the follow-up timelines will be taken in early September 2021, based on progress on recruitment and situation at the time regarding the COVID-19 pandemic.

## Data Availability

This is the study protocol paper.

## Authors’ contributions

**Conceptualization**: Noreen Mdege, Pratima Murthy, Simon Gilbody,

**Funding acquisition:** Najma Siddiqi, Simon Gilbody

**Methodology**: Ian Kellar, Cath Jackson, Heather Thomson, Catherine Hewitt

**Project administration**: Faiza Mujeeb, Krishna Prasad, Gerardo Zavala, Papiya Mazumdar

**Resources**: Najma Siddiqi, Simon Gilbody

**Supervision**: Noreen Mdege, Pratima Murthy, Simon Gilbody

**Writing – original draft:** Gerardo Zavala, Noreen Mdege, Pratima Murthy, Simon Gilbody

**Writing – review & editing:** Gerardo Zavala, Papiya Mazumdar, Noreen Mdege, Pratima Murthy, Simon Gilbody, Faiza Mujeeb, Krishna Prasad, Santosh Kumar Chaturvedi, Arun Kandasamy, Asad Nizami, Baha Ul Haq, Ian Kellar, Cath Jackson, Heather Thomson, David McDaid, Kamran Siddiqi, Catherine Hewitt, Najma Siddiqi

## Acknowledgements

We would like to acknowledge research teams at the National Institute of Mental Health and Neuro-Sciences (NIMHANS) in India and at the Institute of Psychiatry in Pakistan (IOP) for their contribution to the study. We extend our sincere appreciation to Dr. Omara Dogar, Assistant Professor, Department of Health Sciences, University of York for her useful contribution to the intervention mapping work. We want to thank all the participants and family members who would be consenting to participate in the feasibility trial.

## Supporting Information

SPIRIT checklist.

## Appendix 1: Safety consideration

### Research procedures

The researcher will provide assurance on participants anonymity, confidentiality and rights on refusing to answer any uncomfortable question, asking for temporary break or stopping the interview altogether and/ withdrawal from study, without any consequence. It is possible for some questions to cause distress. The researcher will seek help from in-house clinical staff (e.g. counsellors) on participant’s behalf, as and when required.

#### Behavioural support

IMPACT 4S is a low risk interventions. However, in the circumstances of anxiety and distress among participants caused by given intervention information or feedback, researchers will seek help from clinical staff on participant’s behalf.

#### Pharmacological support

Bupropion and nicotine replacement therapies are generally safe medicines with few side effects, and are routinely prescribed for smoking cessation including for persons with severe mental illnesses. Nicotine gum will slowly release small amounts of nicotine in the system of participants, but without the harmful chemicals found in smoked tobacco. The IMPACT 4S participants will be informed about likely side effects from bupropion and nicotine gum and would be advised to make immediate contact with the study team following any experience of side effects. A nearest emergency referral services will be facilitated in case of emergency medical needs. Where the participants would be advised to seek help from, with adequate information provided to the health care providers in regard to their trial participation and current medication usage both within and outside the study. Adherence to medications will be documented in the counsellor’s record book during each session. Standard operating procedures (SoPs) on suicidality would be followed to assess and handle levels of risk severity (e.g., low, medium and high risk) of threats to participant’s own life as expressed through thoughts, feelings or other signs. The participants assessed with *‘Low suicidal risk’* will be recommended for Tele follow up (IVRS-Interactive voice response system) and will be linked with the concerned unit for needed intervention. The participants assessed with *‘Medium and High suicidal risks’* will be advised to get admitted at emergency facility. With prior consents obtained from the participants, family members will be informed about any heightened risks along with appropriate information provided on monitoring and treatment.

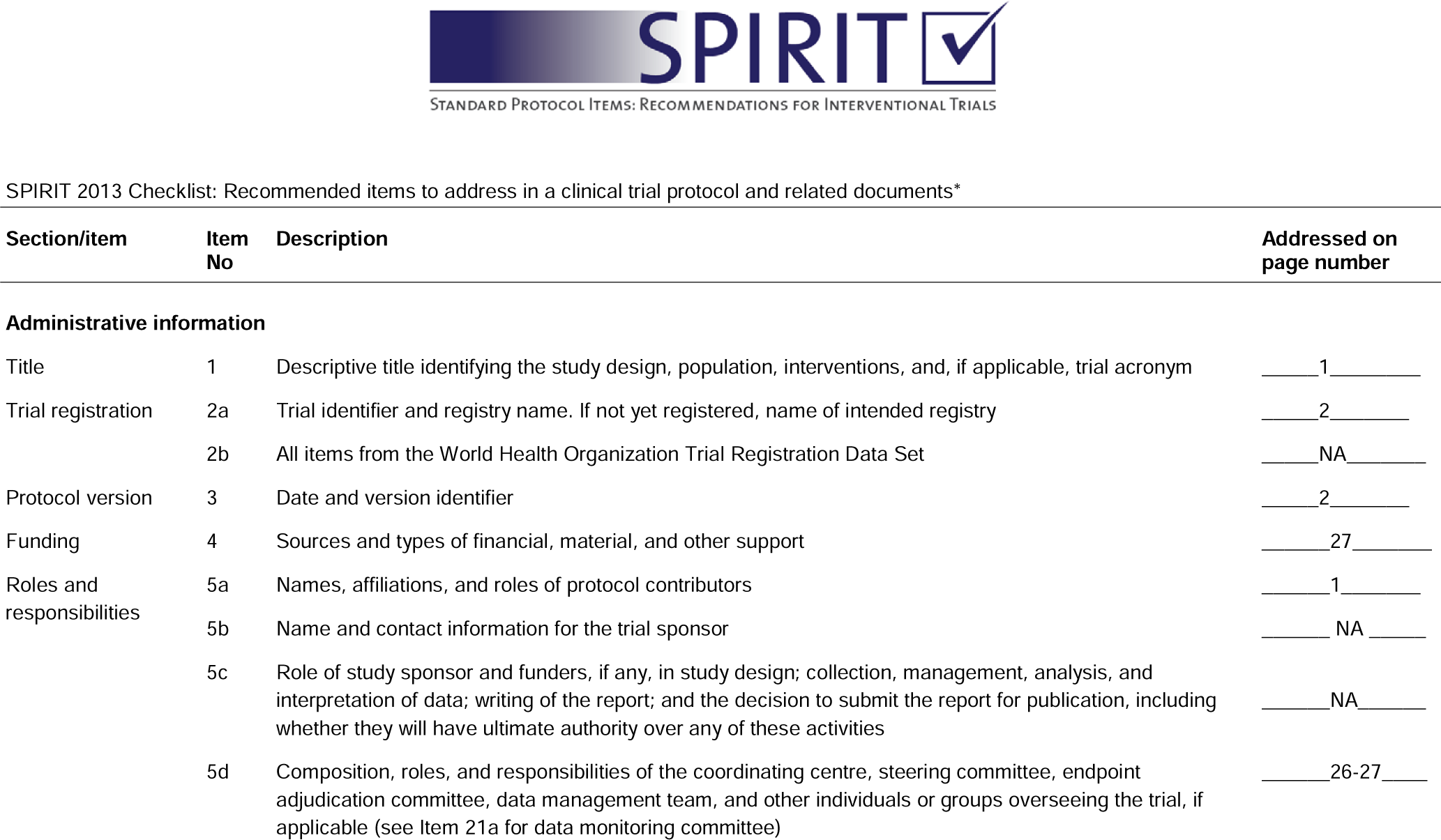
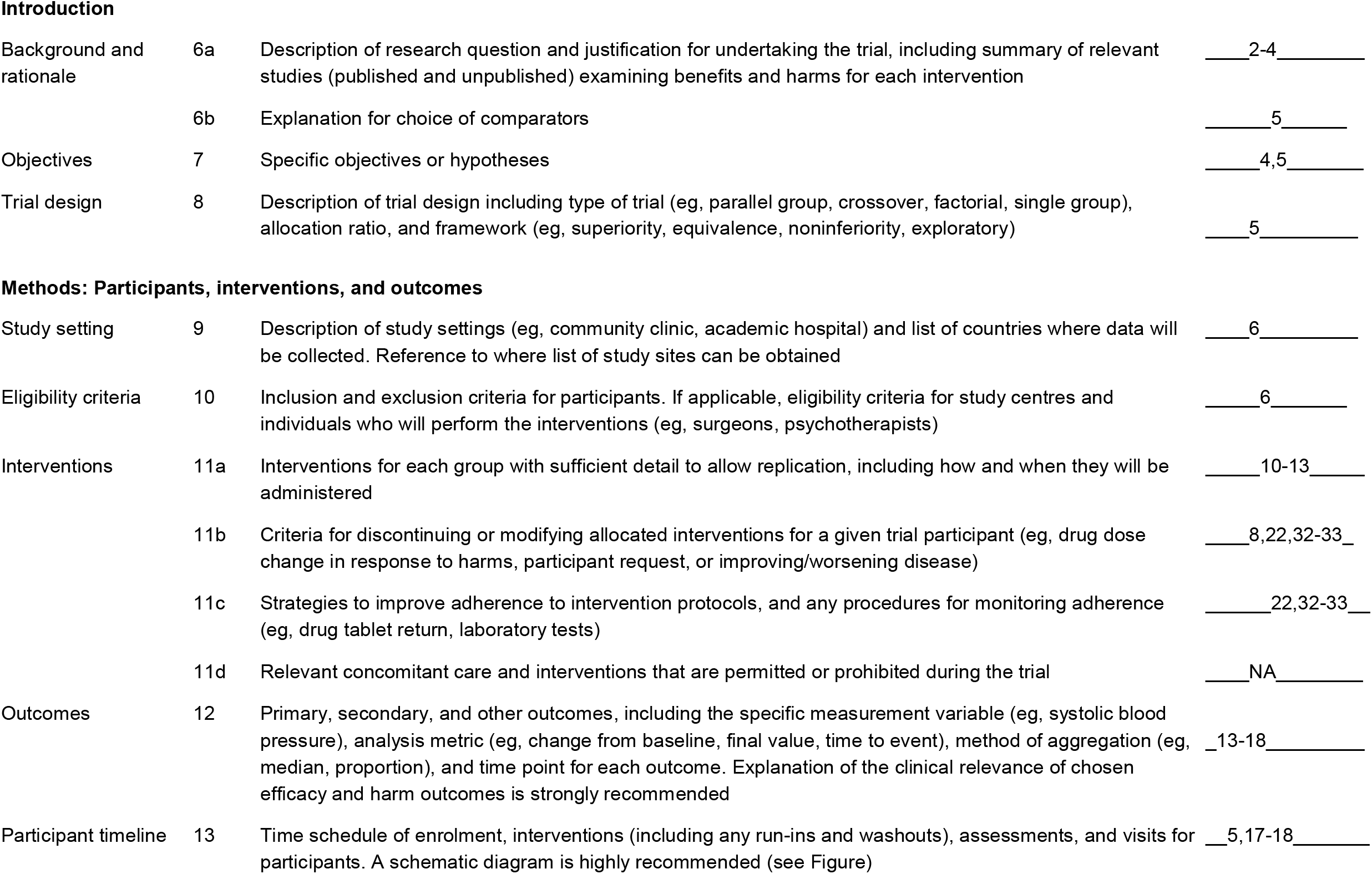
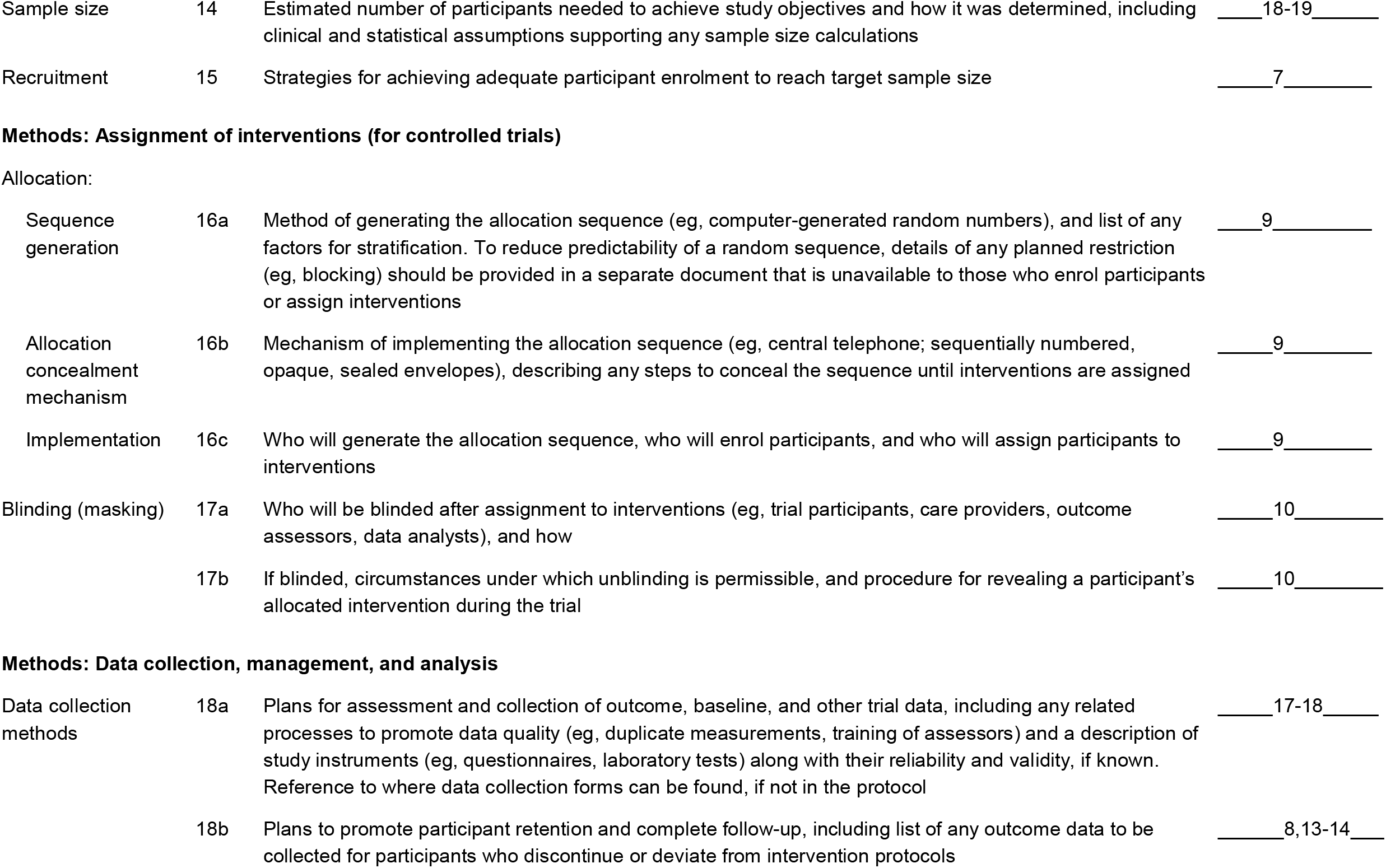
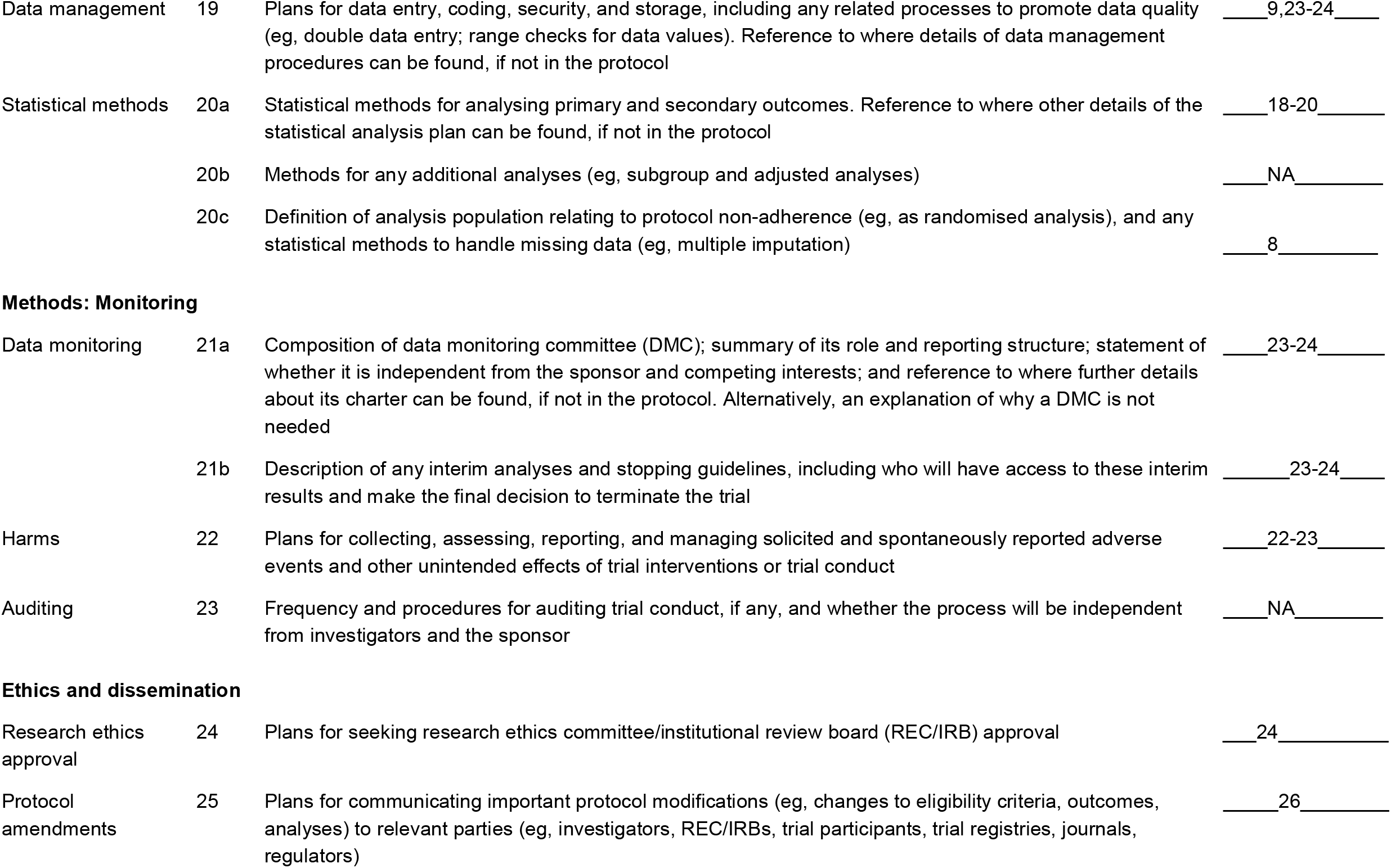
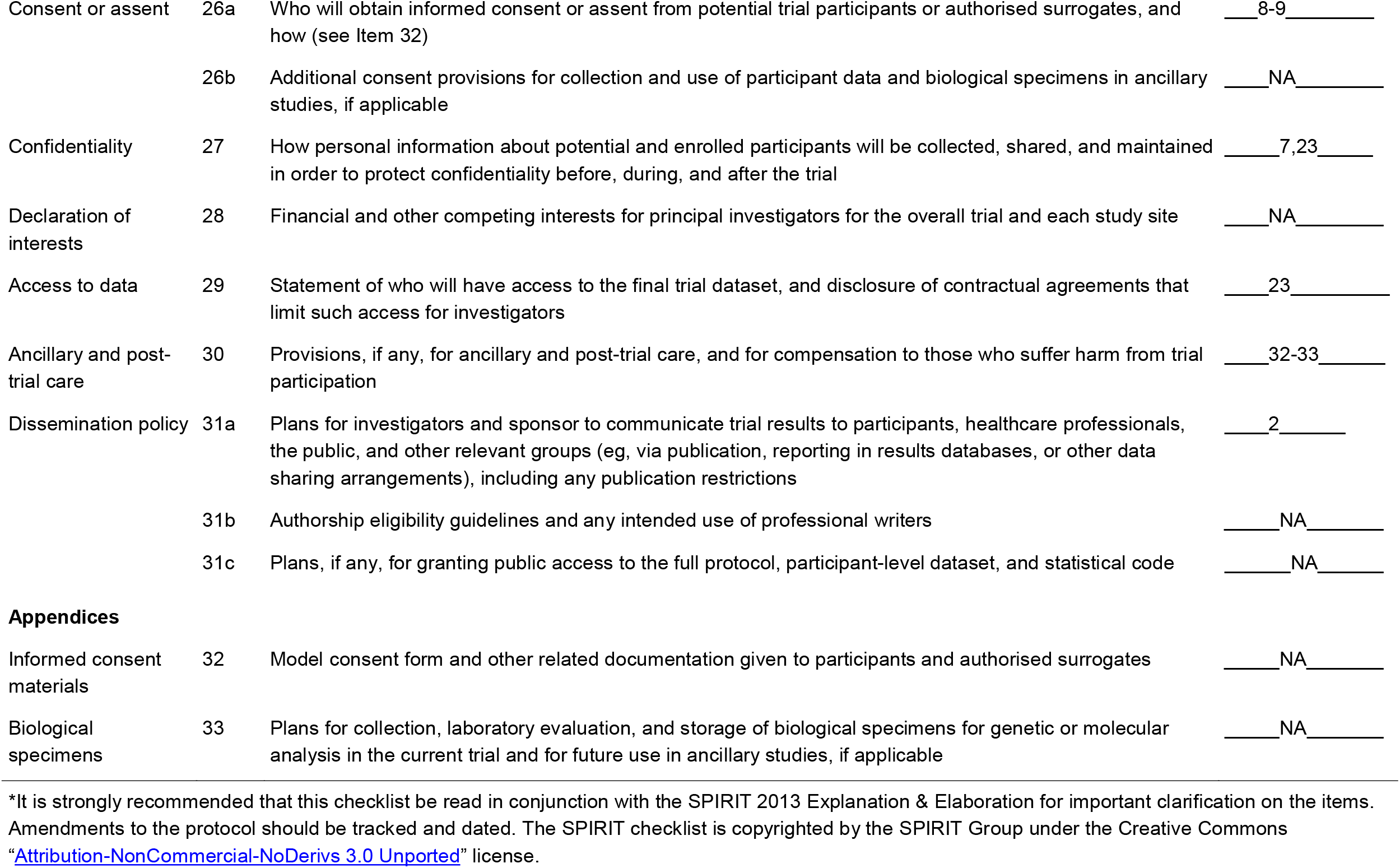

| Section/item | Item No | Description | Addressed on page number |
| --- | --- | --- | --- |
| **Administrative information** |  |  |  |
| Title | 1 | Descriptive title identifying the study design, population, interventions, and, if applicable, trial acronym | ____1____ |
| Trial registration | 2a | Trial identifier and registry name. If not yet registered, name of intended registry | ____2____ |
|  | 2b | All items from the World Health Organization Trial Registration Data Set | ____NA____ |
| Protocol version | 3 | Date and version identifier | ____2____ |
| Funding | 4 | Sources and types of financial, material, and other support | ____27____ |
| Roles and responsibilities | 5a | Names, affiliations, and roles of protocol contributors | ____1____ |
|  | 5b | Name and contact information for the trial sponsor | ____NA____ |
|  | 5c | Role of study sponsor and funders, if any, in study design; collection, management, analysis, and interpretation of data; writing of the report; and the decision to submit the report for publication, including whether they will have ultimate authority over any of these activities | ____NA____ |
|  | 5d | Composition, roles, and responsibilities of the coordinating centre, steering committee, endpoint adjudication committee, data management team, and other individuals or groups overseeing the trial, if applicable (see Item 21a for data monitoring committee) | ____26-27____ |
| Introduction |  |  |  |
| Background and rationale | 6a | Description of research question and justification for undertaking the trial, including summary of relevant studies (published and unpublished) examining benefits and harms for each intervention | ___2-4___ |
|  | 6b | Explanation for choice of comparators | ___5___ |
| Objectives | 7 | Specific objectives or hypotheses | ___4,5___ |
| Trial design | 8 | Description of trial design including type of trial (eg, parallel group, crossover, factorial, single group), allocation ratio, and framework (eg, superiority, equivalence, noninferiority, exploratory) | ___5___ |
| Methods: Participants, interventions, and outcomes |  |  |  |
| Study setting | 9 | Description of study settings (eg, community clinic, academic hospital) and list of countries where data will be collected. Reference to where list of study sites can be obtained | ___6___ |
| Eligibility criteria | 10 | Inclusion and exclusion criteria for participants. If applicable, eligibility criteria for study centres and individuals who will perform the interventions (eg, surgeons, psychotherapists) | ___6___ |
| Interventions | 11a | Interventions for each group with sufficient detail to allow replication, including how and when they will be administered | ___10-13___ |
|  | 11b | Criteria for discontinuing or modifying allocated interventions for a given trial participant (eg, drug dose change in response to harms, participant request, or improving/worsening disease) | ___8,22,32-33___ |
|  | 11c | Strategies to improve adherence to intervention protocols, and any procedures for monitoring adherence (eg, drug tablet return, laboratory tests) | ___22,32-33___ |
|  | 11d | Relevant concomitant care and interventions that are permitted or prohibited during the trial | ___NA___ |
| Outcomes | 12 | Primary, secondary, and other outcomes, including the specific measurement variable (eg, systolic blood pressure), analysis metric (eg, change from baseline, final value, time to event), method of aggregation (eg, median, proportion), and time point for each outcome. Explanation of the clinical relevance of chosen efficacy and harm outcomes is strongly recommended | ___13-18___ |
| Participant timeline | 13 | Time schedule of enrolment, interventions (including any run-ins and washouts), assessments, and visits for participants. A schematic diagram is highly recommended (see Figure) | ___5,17-18___ |
| Sample size | 14 | Estimated number of participants needed to achieve study objectives and how it was determined, including clinical and statistical assumptions supporting any sample size calculations | ____18-19____ |
| Recruitment | 15 | Strategies for achieving adequate participant enrolment to reach target sample size | ____7____ |
| Allocation: |  |  |  |
| Sequence generation | 16a | Method of generating the allocation sequence (eg, computer-generated random numbers), and list of any factors for stratification. To reduce predictability of a random sequence, details of any planned restriction (eg, blocking) should be provided in a separate document that is unavailable to those who enrol participants or assign interventions | ____9____ |
| Allocation concealment mechanism | 16b | Mechanism of implementing the allocation sequence (eg, central telephone; sequentially numbered, opaque, sealed envelopes), describing any steps to conceal the sequence until interventions are assigned | ____9____ |
| Implementation | 16c | Who will generate the allocation sequence, who will enrol participants, and who will assign participants to interventions | ____9____ |
| Blinding (masking) | 17a | Who will be blinded after assignment to interventions (eg, trial participants, care providers, outcome assessors, data analysts), and how | ____10____ |
|  | 17b | If blinded, circumstances under which unblinding is permissible, and procedure for revealing a participant's allocated intervention during the trial | ____10____ |
| **Methods: Data collection, management, and analysis** |  |  |  |
| Data collection methods | 18a | Plans for assessment and collection of outcome, baseline, and other trial data, including any related processes to promote data quality (eg, duplicate measurements, training of assessors) and a description of study instruments (eg, questionnaires, laboratory tests) along with their reliability and validity, if known. Reference to where data collection forms can be found, if not in the protocol | ____17-18____ |
|  | 18b | Plans to promote participant retention and complete follow-up, including list of any outcome data to be collected for participants who discontinue or deviate from intervention protocols | ____8,13-14____ |
| Data management | 19 | Plans for data entry, coding, security, and storage, including any related processes to promote data quality (eg, double data entry; range checks for data values). Reference to where details of data management procedures can be found, if not in the protocol | ____9,23-24____ |
| Statistical methods | 20a | Statistical methods for analysing primary and secondary outcomes. Reference to where other details of the statistical analysis plan can be found, if not in the protocol | ____18-20____ |
|  | 20b | Methods for any additional analyses (eg, subgroup and adjusted analyses) | ____NA____ |
|  | 20c | Definition of analysis population relating to protocol non-adherence (eg, as randomised analysis), and any statistical methods to handle missing data (eg, multiple imputation) | ____8____ |
| **Methods: Monitoring** |  |  |  |
| Data monitoring | 21a | Composition of data monitoring committee (DMC); summary of its role and reporting structure; statement of whether it is independent from the sponsor and competing interests; and reference to where further details about its charter can be found, if not in the protocol. Alternatively, an explanation of why a DMC is not needed | ____23-24____ |
|  | 21b | Description of any interim analyses and stopping guidelines, including who will have access to these interim results and make the final decision to terminate the trial | ____23-24____ |
| Harms | 22 | Plans for collecting, assessing, reporting, and managing solicited and spontaneously reported adverse events and other unintended effects of trial interventions or trial conduct | ____22-23____ |
| Auditing | 23 | Frequency and procedures for auditing trial conduct, if any, and whether the process will be independent from investigators and the sponsor | ____NA____ |
| **Ethics and dissemination** |  |  |  |
| Research ethics approval | 24 | Plans for seeking research ethics committee/institutional review board (REC/IRB) approval | ____24____ |
| Protocol amendments | 25 | Plans for communicating important protocol modifications (eg, changes to eligibility criteria, outcomes, analyses) to relevant parties (eg, investigators, REC/IRBs, trial participants, trial registries, journals, regulators) | ____26____ |
| Consent or assent | 26a | Who will obtain informed consent or assent from potential trial participants or authorised surrogates, and how (see Item 32) | ___8-9___ |
|  | 26b | Additional consent provisions for collection and use of participant data and biological specimens in ancillary studies, if applicable | ___NA___ |
| Confidentiality | 27 | How personal information about potential and enrolled participants will be collected, shared, and maintained in order to protect confidentiality before, during, and after the trial | ___7,23___ |
| Declaration of interests | 28 | Financial and other competing interests for principal investigators for the overall trial and each study site | ___NA___ |
| Access to data | 29 | Statement of who will have access to the final trial dataset, and disclosure of contractual agreements that limit such access for investigators | ___23___ |
| Ancillary and post-trial care | 30 | Provisions, if any, for ancillary and post-trial care, and for compensation to those who suffer harm from trial participation | ___32-33___ |
| Dissemination policy | 31a | Plans for investigators and sponsor to communicate trial results to participants, healthcare professionals, the public, and other relevant groups (eg, via publication, reporting in results databases, or other data sharing arrangements), including any publication restrictions | ___2___ |
|  | 31b | Authorship eligibility guidelines and any intended use of professional writers | ___NA___ |
|  | 31c | Plans, if any, for granting public access to the full protocol, participant-level dataset, and statistical code | ___NA___ |
| Informed consent materials | 32 | Model consent form and other related documentation given to participants and authorised surrogates | ___NA___ |
| Biological specimens | 33 | Plans for collection, laboratory evaluation, and storage of biological specimens for genetic or molecular analysis in the current trial and for future use in ancillary studies, if applicable | ___NA___ |

\*It is strongly recommended that this checklist be read in conjunction with the SPIRIT 2013 Explanation & Elaboration for important clarification on the items. Amendments to the protocol should be tracked and dated. The SPIRIT checklist is copyrighted by the SPIRIT Group under the Creative Commons "Attribution-NonCommercial-NoDerivs 3.0 Unported" license.

